# The risk of SARS-CoV-2 Omicron variant emergence in low and middle-income countries (LMICs)

**DOI:** 10.1101/2022.01.14.22268821

**Authors:** Kaiming Bi, Jose Luis Herrera-Diestra, Yuan Bai, Zhanwei Du, Lin Wang, Graham Gibson, Maureen Johnson-Leon, Spencer J. Fox, Lauren Ancel Meyers

## Abstract

We estimated the probability of undetected emergence of the SARS-CoV-2 Omicron variant in 25 low and middle-income countries (LMICs) prior to December 5, 2021. In nine countries, the risk exceeds 50%; in Turkey, Pakistan and the Philippines, it exceeds 99%. Risks are generally lower in the Americas than Europe or Asia.

## Main text

The B.1.1.529 SARS-COV-2 variant was first detected and reported in South Africa on November 26^th^, 2021 [1]. By December 5^th^, 2021, more than 40 countries reported Omicron variant cases. Most of these countries are developed and high-income countries, including the United Kingdom, the United States, and Netherlands [2]. However, low and middle income countries (LMICs) may be less likely to detect a new variant [3] and more vulnerable to catastrophic public health outcomes than high-income countries, because of lower capacity for COVID-19 testing, vaccination, and medical treatment [4, 5]. Only a few LMICs have direct flights from the countries in Southern Africa where Omicron was initially detected [6] and many enacted border policies to reduce Omicron importation risks from these countries [7]. However, LMICs were at risk for Omicron importations from large international destinations outside of Southern Africa in which Omicron emerged in late 2021.

We analyzed the risks of the Omicron variant importation in 25 LMICs in which Omicron was not reported as of December 5, 2021: Bangladesh, Nepal, Philippines, Colombia, Egypt, Pakistan, Paraguay, Turkey, Serbia, Bolivia, Argentina, Uruguay, Bhutan, Indonesia, Albania, Jordan, Panama, Dominican Republic, Ecuador, Peru, Jamaica, Honduras, Guatemala, Costa Rica, and El Salvador. We first estimated the daily travel volume to each country from 13 large countries in which Omicron had already been detected, based on data from Facebook Data for Good (Figure A) [8]. We estimated the prevalence of Omicron in each of the 13 Omicron detected countries (ODCs) assuming that only 2.5% of early cases were identified and reported [2, 9], and then estimated the probability of travel-based introductions into each LMIC by December 5, 2021 (see Supporting Information). The European LMICs (Serbia and Turkey), which are highly connected to Western European countries that reported Omicron cases by November 2021, have the highest estimated risks, followed by the Asian LMICs (Pakistan, Bangladesh, and Nepal), with high inflows of travelers from South Africa (via connecting flights), the UK, and India. LMICs in the Americas (Colombia, Dominican Republic, and Paraguay) are primarily at risk for importations from the US and Brazil. We estimate that 6 of the 25 studied LMICs had over a 50% chance of having received at least one travel-based Omicron importation from ODCs by December 5, 2021 (Figure B).

**Figure:**
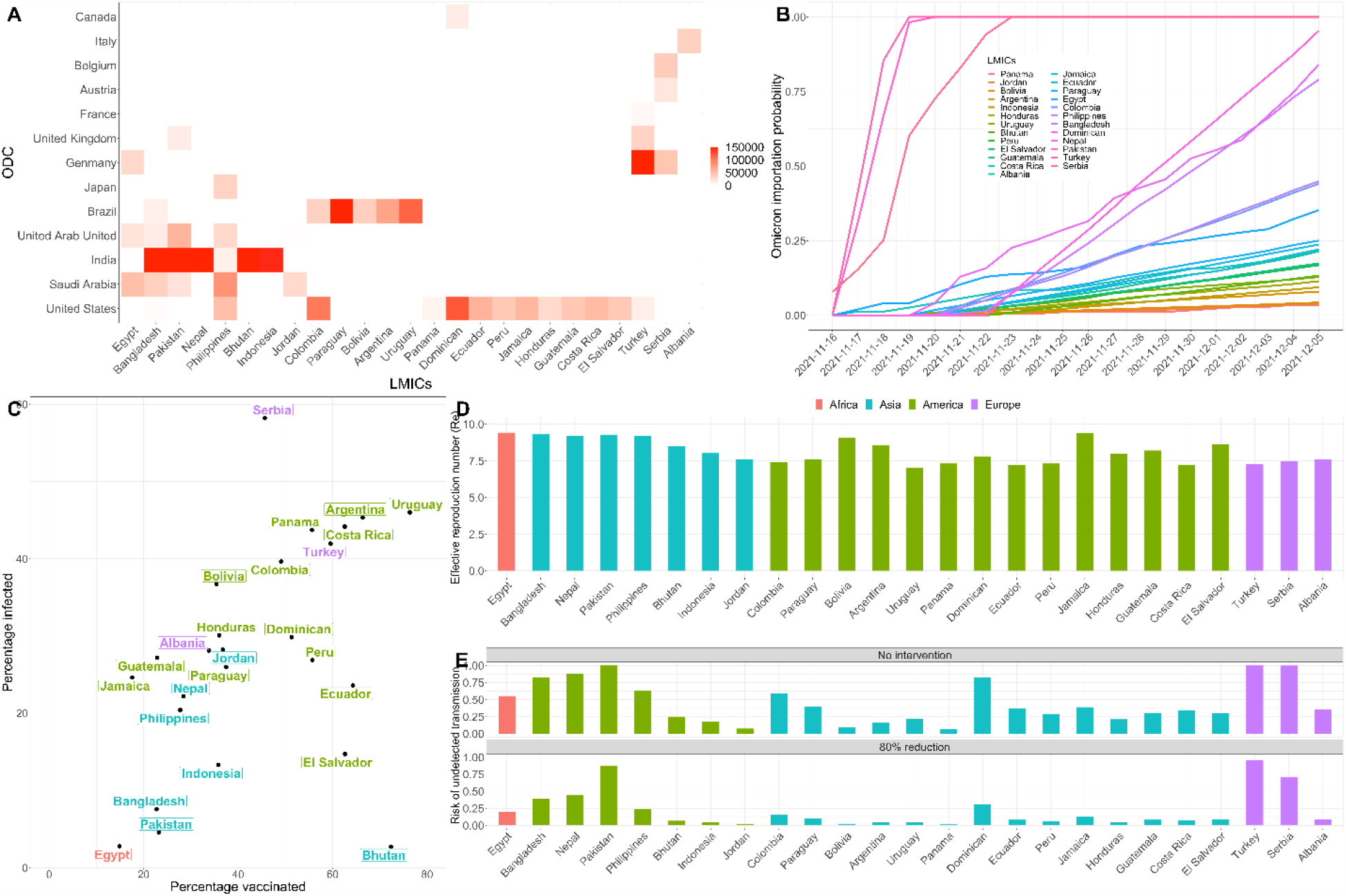
Risks of Omicron emergence in 25 low- and middle-income countries by December 5, 2021. A. Cumulative number of travelers to each of the 25 LMICs reported by Facebook Data for Good from November 16 to December 5, 2021 from 13 countries in which Omicron had already been detected by December 5 (ODCs): Belgium, Italy, United Kingdom, Germany, Canada, Austria, Japan, Brazil, United State, United Arab Emirates, Saudi Arabia, France, India [8]. B. The probability of receiving at least one Omicron importation via travelers from the 13 ODCs between November 16 and December 5, 2021. C. Estimated vaccination and prior infection rates for each LMIC, assuming a 40% infection reporting rate for all countries [15]. D. Estimated effective reproduction number for the Omicron variant in each LMIC, assuming that Omicron is twice as transmissible as the Delta variant and that vaccines have 50% lower efficacy against Omicron in comparison to Delta. E. Comparisons of the estimated risks of the undetected Omicron transmission in LMICs with and without intervention that reduces transmission by 80%.

To assess the risk of Omicron transmission following importation, we estimate the immunity-based effective reproduction number (*R*_*e*_) in each LMIC as of December 5, 2021, based on reported vaccination levels [10], estimates of infection-acquired immunity [11] (Figure C), and recent estimates for the transmissibility and immune-evasiveness of the Omicron variant [12] (see Supporting Information). Recent studies have estimated that Omicron has double the reproduction number of the Delta variant, suggesting a basic reproduction number (*R*_0_) of 11.88 (95%CI: 9.16-14.61) (Figure SI.3) [13]. The same study also suggests that SARS-CoV-2 vaccines are 50% less effective (VE) against Omicron compared to Delta (Figure SI.5) [13]. Given the low vaccination coverage and high infectivity of Omicron, we estimate that the immunity-based *R*_*e*_ of Omicron on December 5, 2021 ranges from 7.0 to 9.4 across the 25 studied LMICs, without additional public health interventions (Figure D).

Combining our estimates of Omicron importation and transmission risks, we estimate the probability of undetected Omicron transmission in LMICs by early December (Figure E-details see Supporting Information). Among the studied LMICs, Turkey (99.99%), Pakistan (99.95%), and Serbia (99.81%) have the highest estimated risk, followed by Nepal (87.98%), Bangladesh (84.86%), and the Dominican Republic (82.21%). If these countries implement non-pharmaceutical interventions that reduce transmission by 80%, the probability of undetected emergence declines by 12.02% to 80.77%% across the 25 LMICs (Figure E). Given the high socioeconomic costs of travel restrictions and some non-pharmaceutical interventions, many of these LMICs did not take measures to prevent introductions or slow spread [14]. Our analyses suggest that SARS-CoV-2 variants like Omicron can rapidly emerge in LMICs and spread for weeks before detection.

## Supporting information

https://docs.google.com/document/d/1acm04iY2SxE0g8d-MlsQuIejcRl_NRvH/edit?rtpof=true

## Data Availability

All data produced in the present study are available upon reasonable request to the authors

## Acknowledgements

This research was supported by grants from the US National Institutes of Health (grant no. R01 AI151176) and the US Centers for Disease Control and Prevention (grant no. U01 IP001136) and a donation from Love, Tito’s (the philanthropic arm of Tito’s Homemade Vodka, Austin, TX, USA) to the University of Texas to support the modeling of COVID-19 mitigation strategies.

## Ethics Declaration

The authors were granted by Facebook Data for Good team to use the Travel Pattern data in research. The results of the research conducted are approved for sharing. The access to the Facebook Data for Good database for research purpose could be granted after the registration.

## Reference

[1] WHO. 2021. “Update on Omicron.” Www.who.int. November 28, 2021. https://www.who.int/news/item/28-11-2021-update-on-omicron.

[2] “GISAID - HCov19 Variants.” n.d. Www.gisaid.org. https://www.gisaid.org/hcov19-variants/.

[3] Sisa, Ivan, Marco Fornasini, and Enrique Teran. “COVID-19 research in LMICs.” The Lancet 398, no. 10307 (2021): 1212–1213.

[4] Helmy, Mohamed, Mohamed Awad, and Kareem A. Mosa. “Limited resources of genome sequencing in developing countries: challenges and solutions.” Applied & translational genomics 9 (2016): 15–19.

[5] Siow, Wen Ting, Mei Fong Liew, Babu Raja Shrestha, Faisal Muchtar, and Kay Choong See. “Managing COVID-19 in resource-limited settings: critical care considerations.” Critical Care 24, no. 1 (2020): 1–5.

[6] Flightradar24. n.d. “Live Flight Tracker - Real-Time Flight Tracker Map.” Flightradar24. Accessed December 12, 2021. https://www.flightradar24.com/data/airports.

[7] Mallapaty, Smriti. “Omicron-variant border bans ignore the evidence, say scientists.” Nature.

[8] “Data for Good Tools and Data.” n.d. http://Dataforgood.facebook.com. https://dataforgood.facebook.com/dfg/tools.

[9] : Davis, Jessica T., Matteo Chinazzi, Nicola Perra, Kunpeng Mu, Marco Ajelli, Natalie E. Dean, Corrado Gioannini et al. “Cryptic transmission of SARS-CoV-2 and the first COVID-19 wave.” Nature (2021): 1–9.

[10] Mathieu, E., Ritchie, H., Ortiz-Ospina, E. et al. A global database of COVID-19 vaccinations. Nat Hum Behav (2021). https://doi.org/10.1038/s41562-021-01122-8

[11] World Health Organization. 2021. “WHO COVID-19 Dashboard.” Covid19.Who.int. World Health Organization. 2021. https://covid19.who.int/.

[12] Miller, Joel C., Anja C. Slim, and Erik M. Volz. “Edge-based compartmental modelling for infectious disease spread.” Journal of the Royal Society Interface 9, no. 70 (2012): 890–906.

[13] Chen, Jiahui, Rui Wang, Nancy Benovich Gilby, and Guo-Wei Wei. “Omicron (B. 1.1. 529): Infectivity, vaccine breakthrough, and antibody resistance.” arXiv preprint 2112.01318 (2021).

[14] Torres-Rueda, Sergio, Sedona Sweeney, Fiammetta Bozzani, Nichola R. Naylor, Tim Baker, Carl Pearson, Rosalind Eggo et al. “Stark choices: exploring health sector costs of policy responses to COVID-19 in low-income and middle-income countries.” BMJ global health 6, no. 12 (2021): e005759.

[15] Irons, Nicholas J., and Adrian E. Raftery. “Estimating SARS-CoV-2 Infections from Deaths, Confirmed Cases, Tests, and Random Surveys.” arXiv preprint 2102.10741 (2021).

