## Supplementary material for "The risk of SARS-CoV-2 Omicron variant emergence in low and middle-income countries (LMICs)": https://docs.google.com/document/d/1acm04iY2SxE0g8d-MlsQuIejcRl_NRvH/edit?rtpof=true

**The Facebook user mobility data and estimated traveler population**

Facebook Data for good team collected the travel patterns, which show the number of Facebook users moving among countries via air, train, or car [1]. We focus on the Facebook travelers departing from Omicron detected counties (ODCs) to LMICs by December 5th, 2021. We assumed that the actual Omicron introduction took place 10 days before Omicron was first reported in the studied ODCs [2, 4] (as shown in Table SI.1). The patterns of Facebook mobility data from November 16, 2021 to December 5, 2021 are shown in Fig SI.1. (a) and (b).

**Table SI.1. Date of Omicron detection and potential introduction for the studied ODCs**

| Studied ODCs | Date of Omicron detection | Estimated date of Omicron introduction |
|---|---|---|
| Belgium | November 26,2021 | November 16,2021 |
| Neitherland | November 26,2021 | November 16,2021 |
| United Kingdom | November 27, 2021 | November 17,2021 |
| Germany | November 27, 2021 | November 17,2021 |
| Italy | November 27, 2021 | November 17,2021 |
| Canada | November 28, 2021 | November 18,2021 |
| Austria | November 29, 2021 | November 19,2021 |
| Brazil | November 30, 2021 | November 20, 2021 |
| Japan | November 30, 2021 | November 20, 2021 |
| United State | December 1, 2021 | November 21, 2021 |
| Saudi Arabia | December 1, 2021 | November 21, 2021 |
| United Arab Emirates | December 1, 2021 | November 21, 2021 |
| France | December 2, 2021 | November 22, 2021 |
| India | December 3, 2021 | November 23, 2021 |

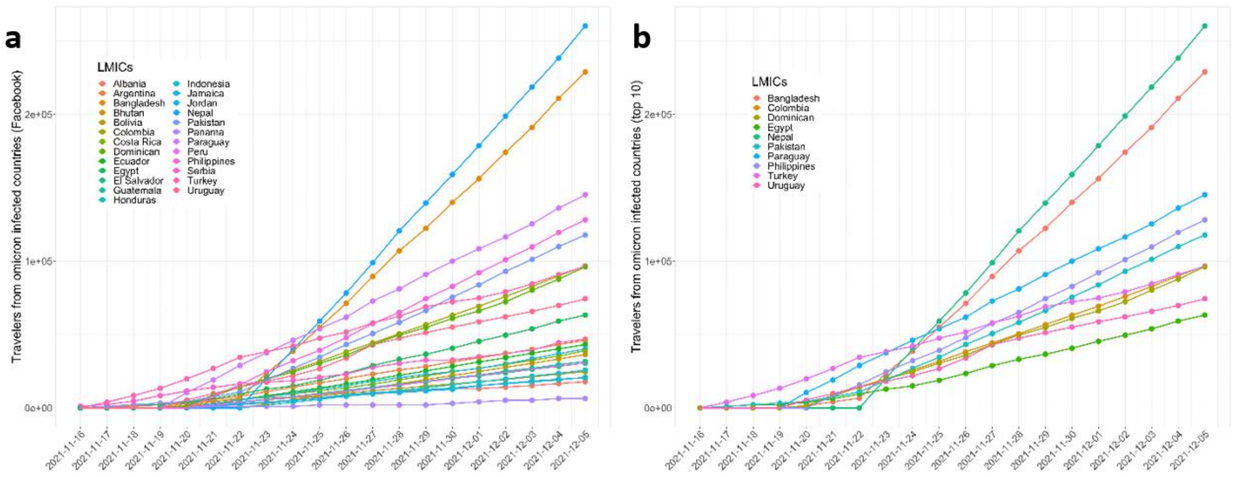


**Figure SI. 1. Facebook international travelers arriving at the LMICs by December 5, 2021.** *(a) Summarized data for all studied LMICs, which have more than 1,000 Facebook travelers in a day; (b) Summarized data for top 10 LMICs by the number of Facebook travelers.*

Since international travelers include both Facebook users and non-Facebook users, we adjust to compensate for non-Facebook users. Based on the ratios of Facebook users for the studied countries [3], we estimated the total number of travelers departing from ODCs to LMICs by December 5th, 2021 as

$$LMIC_t^{total} = LMIC_t^{Facebook} * \left(\frac{Facebook\ users_{LMIC}}{Population_{LMIC}}\right), \quad \text{(SI.1)}$$

where $LMIC_t^{total}$ and $LMIC_t^{Facebook}$ represent the number of travelers and Facebook users departing from ODCs to the selected LMIC at day $t$, respectively. $Facebook\ users_{LMIC}$ and $Population_{LMIC}$ are the total numbers of Facebook users and population at the selected LMIC in the time period considered. The estimated values of the international travelers for each LIMC are shown in Fig SI. 2. (a) and (b).

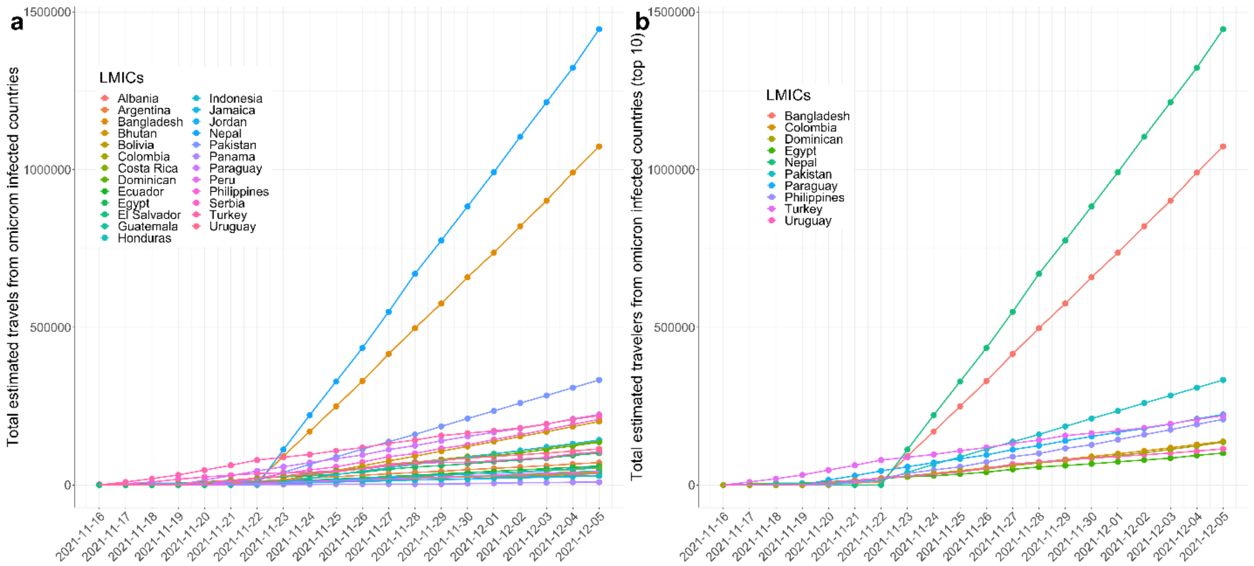


**Figure SI. 2. International travelers departing from the ODCs by December 5, 2021.** *(a) Summarized data for all studied LMICs; (b) Summarized data for top 10 LMICs by the number of international travelers.*

**The estimation of the Omicron importation risk based on the prevalence data**

We estimated the risk that the Omicron was introduced into a LMIC via travelers from ODCs, by multiplying the probability that one traveler is an Omicron carrier with the number of international travelers leaving from an ODC to the studied LMIC. The prevalence data for the ODC is estimated based on the Omicron reported cases [4]. Since previous studies showed that only 1-4% of the COVID-19 cases were actually reported at the early stage of the epidemic, we assumed that only 2.5% of the Omicron cases were reported by December 5, 2021.

$$N_{ODC\to LMIC}(t) = ODC \to LMIC_t^{total} * \left(\frac{Prevalence_{ODC}}{Population_{ODC}}\right) \quad \text{(SI.2)}$$

$$N_{LMIC}(t) = \sum_{ODCs} \quad N_{ODC\to LMIC}(t) \quad \text{(SI.3)}$$

Where $N_{ODC\to LMIC}(t)$ is the estimated Omicron cases imported from a selected ODC to a selected LMIC at day t; $N_{LMIC}(t)$ is the summarized Omicron cases imported for the selected LMIC at day t; $ODC \to LMIC_t^{total}$ is the the number of travelers departing from a specific ODC to the selected LMIC at day $t$; $Prevalence_{ODC}$ and $Population_{ODC}$ are the total numbers of the estimated Omicron cases and the population at the selected ODC (detailed shown in Table SI.9). Therefore, the $\left(\frac{Prevalence_{ODC}}{Population_{ODC}}\right)$ represents the estimated possibility that a travel (traveling from ODC to LMIC) is Omicron infected.

**The calculation of $R_e$ and the estimation of the Omicron spreading risk**

The basic reproduction number ($R_0$) has been calculated for the SIR-like edge-based compartmental model [5]. In this order of ideas, in a completely naive population, $R_0$ of COVID-19 can be calculated as:

$$R_0 = \frac{\beta}{\beta+\gamma}\frac{<k^2-k>}{<k>} \quad \text{(SI.4)}$$

Where $\beta$ is the infection rate for the naïve population, $\gamma$ is the recovery rate, $< k >$ and $< k^2 >$ are the mean degree and mean of the squared degree for the studied network. Due to the lack of degree distribution data for the studied LMICs, we assumed the population distribution in LMICs is similar to the rural population distribution for the developed countries [30]. Therefore, we assumed the population network in LMICs follows the exponential distribution with $< k >= 10$ and $< k^2 >$=184.

If the studied population is not completely naïve (partially protected by nature immunity and vaccination), the effective reproduction number is calculated as the transmissivity. In this context, we use $\beta'$ to represent the average infection rate for a partially immunized population.

$$R_e = \frac{\beta'}{\beta'+\gamma}\frac{< k^2 - k >}{< k >}$$

$$= \frac{(\beta(1-imm)+\beta(1-\alpha)imm)}{(\beta(1-imm)+\beta(1-\alpha)imm)+\gamma}\frac{<k^2-k>}{<k>}$$

$$= \frac{\beta(1-\alpha*imm)}{\beta(1-\alpha*imm)+\gamma}\frac{<k^2-k>}{<k>} \quad \text{(SI.5)}$$

Where $\alpha$ is the efficiency of the immunity, the portions of the immunized population and naïve population are $imm$ and $(1 - imm)$ respectively. The immunized population is defined as the population with immunity from previous infections and the uninfected population with vaccinations.

$$imm = Prevalence\% + (1 - Prevalence\%) * vac\% \quad \text{(SI.6)}$$

Assumed the studied population is homogeneous , the probability that Omicron will not spread is $(1/R_e)^N$, where $N$ is the cumulative number of imported Omicron cases for the LMIC from November 16 to December 5, 2021.. Based on that assumption , we estimated the risk of Omicron spreading in this LMIC ($Risk_{LMIC-sp}$) as:

$$Risk_{LMIC-sp} = 1 - (1/R_e)^N \quad \text{(SI.6)}$$

**Literature review of the observed $R_0$ for delta and estimation of $R_0$ for Omicron**

There are no existing literature directly reporting the $R_0$ of the Omicron variant. The most recent work shows that the Omicron variant may be about twice as infectious as the Delta variant [6]. Therefore, we first review the literature in the $R_0$ of Delta variant, then estimate the $R_0$ of Omicron variant as two times the $R_0$ of Delta variant.

**Table SI.2. Literature review of the observed $R_0$ for the "delta variant"**

| Where & when | Estimated R0 | Resources |
|---|---|---|
| Guangdong (China), May-Jun 2021 | 3.2 (2-5) | Zhang et.al. [7]<br>Shi et.al. [8] |
| India, May 2021 | Higher than 4.5 | Mandal [14] |
| England, before 9 Jun 2021 | (5-8) | SPI-M-O [9]<br>David et.al. [10] |
| New Zealand, 2021 | (4.5-6) | Nguyen [15] |
| US California, by summer, 2021 | (4.6-5.0) | Head [12] |
| China general, July, 2021 | 50% higher than the alpha variant | Liu et.al. [11] |
| Demark, 17 May to 25 July, 2021 | 3.28 times the original strain | Hansena [13] |
| France, June, 2021 | 79% greater than the alpha variant (52-110%) | Alizon [16] |
| Global, by 3 Jun, 2021 | 55% greater than the alpha variant (43-68%) | Campbell [17] |

Based on the literature review, we know the $R_0$ of the Delta variant is 5.94 (95%CI: 4.58-7.30). Therefore, the $R_0$ of the Omicron variant is estimated as 11.88 (95%CI: 9.16-14.61). Meaning, the transmission ability of the Omicron variant is extremely high in a completely naive population, which has no immunity and public health intervention to protect themselves.

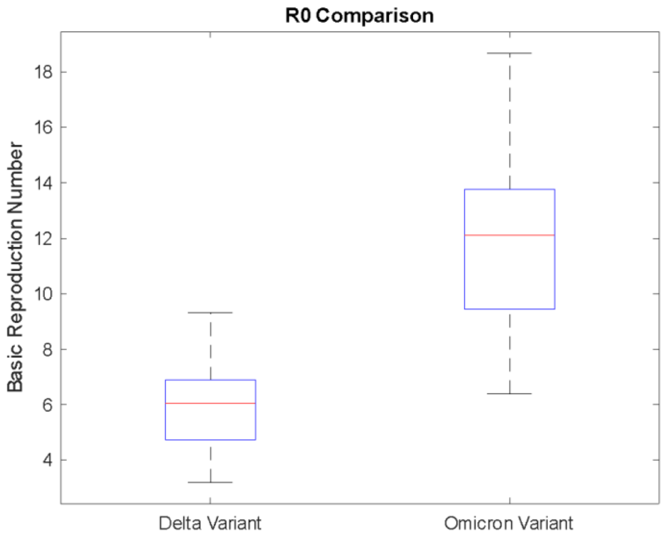


**Figure SI.3. Observed $R_0$ Comparisons for "Delta variant" and "Omicron variant"**

**The estimation of $\gamma$ and $\beta$ of Omicron (contributed in the calculation for $R_e$ of Omicron)**

We did another literature review to estimate the recovery rate $\gamma$. The details of the literature are summarized in Table SI.3. Based on these data, we estimated the recovery rate $\gamma$ is 0.0756 per day (95%CI: 0.05-0.10), as shown in Table SI.4 and Figure SI.4.

**Table SI.3. Literature review of the recovery rate $\gamma$**

| When and where | Observed $\gamma$ value | Resources |
|---|---|---|
| Wuhan China, Jan to Mar 2020 | 0.01-0.03 | Zareie [15] |
| 13 Counties, by Aug 2020 | 1/(7-14 days) 0.107 | Walsh [16] |
| Australian, by July 2020 | 1/(10-18 days) 0.078 | Basile [17] |
| Canada, by Aug 2020 | 1/(8 days) 0.125 | Bullard [18] |
| Germany, by Jun 2020 | 1/(18-21 days) 0.05 | Decker [19] |
| Spain, by Jun 2020 | 1/(10-32 days) 0.066 | Folgueira [20] |
| Korea, Jan to Mar 2020 | 1/(11-15 days) 0.079 | Jeong [21] |
| United State, by Mar 2020 | 1/(8 days) 0.125 | Kujawski [22] |
| Taiwan, by April 2020 | 1/(18 days) 0.056 | Zhang [14] |

| | | |
|---|---|---|
| Netherlands, Mar to Apr 2020 | 1/(20 days) 0.05 | Van Kampen [23] |

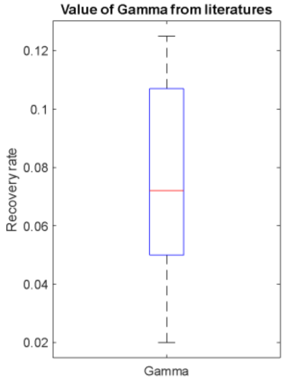


**Figure SI.4. Observed recovery rate $\gamma$**

**Table SI.4. Statistics of the recovery rate $\gamma$**

| | $\gamma$ value | Recover time |
|---|---|---|
| Mean | 0.0756 | 13.2 days |
| Median | 0.0720 | 13.9 days |
| 95% CI | 0.05-0.10 | 10-20 days |

Based on equation (SI.5), $\beta$ can be calculated based on the observation of $R_0, \gamma$ and $< k >$. Assume the network follows the exponential distribution with $< k >= 10$, then $\frac{<k^2-k>}{<k>}$ is equal to 17.4. Then we estimated the $\beta$ for "omicron variant" with $< k >= 10$ as:

$$\beta_{Omicron} = \frac{R_0\gamma}{(\frac{<k^2-k>}{<k>} - R_0)} = 0.1593 \quad \text{(SI.7)}$$

**Literature review of the vaccine efficacy (VE) for delta and estimation of VE for Omicron**

There is no existing literature directly reporting the VE of the Omicron variant. The most recent work shows that Omicron may be twice more likely to escape current vaccines than the Delta variant [6]. Therefore, we first review the literature in the VE for Pfizer (BnT162b2), Moderna (mRnA-1273), J&J (Ad26.CoV2-S), and AZ (AZD1222) vaccines to Delta variant (Table SI.5-8), then estimate the VE of Omicron variant as 50% of the VE of Delta variant. Since the vaccination time varies greatly in different studied LMICs, we didn't consider the VE incorporated with the waning immunity.

**Table SI.5. Observed VE of Pfizer BnT162b2**

| Variant & when | Estimated VE | Resources |
|---|---|---|
| Delta, UK, May 2021 | 79% (75%-82%) | Sheikh et.al [18] |
| Delta, UK, May 2021 | 88% (85.3-90.1%) | Bernal et.al [19] |
| Delta, Canada, June 2021 | 87% (64%-95%) | Nasreen et. al [20] |
| Delta, Qatar, July 2021 | 53.5%(43.9%-61.4%) | Tang et.al [21] |
| Delta, US, August 2021 | 83.4% (74% - 89.4% | Bajema et.al [22] |

**Table SI.6. Observed VE of Moderna mRnA-1273**

| Variant & when | Estimated VE | Resources |
|---|---|---|
| Delta, Canada, June 2021 | 72% | Nasreen et. al [20] |
| Delta, Qatar, July 2021 | 84.8% (75.9%-90.8%) | Tang et.al [21] |
| Delta, US, August 2021 | 91.6% (83.5% - 95.7% | Bajema et.al [22] |
| Delta, US, after July 2021 | 76% (58% - 87%) | Puranik et.al [23] |
| Delta, US, March 2021 | 48.4% (42.3% - 53.8%) | Nanduri et.al [25] |

| | | |
|---|---|---|
| Delta, US, after July 2021 | 42% (13% - 62%) | Puranik et.al [23] |
| Delta, US, August 2021 | 52.2% (47.7% - 56.3%) | Nanduri et.al [25] |
| Delta, US, July 2021 | 74% (65% – 82%) | Barlow et.al [27] |

| | | |
|---|---|---|
| Delta, US, July 2021 | 74% (65% – 82%) | Barlow et.al [27] |

**Table SI.7. Observed VE of J&J Ad26.CoV2-S**

| Variant & when | Estimated VE | Resources |
|---|---|---|
| Delta, June, 2021 | 60% | CBS News [24] |
| Delta, US, July 2021 | 77% (74% - 79%) | Polinski et.al [26] |
| Delta, US, July 2021 | 51% (0 – 76%) | Barlow et.al [27] |

**Table SI.8. Observed VE of AZ AZD1222**

| Variant & when | Estimated VE | Resources |
|---|---|---|
| Delta, UK, May 2021 | 67% (61.3-71.8%) | Bernal et.al [19] |
| Delta, Canada, June 2021 | 61% | Nasreen et. al [20] |
| Delta, June, 2021 | 60% | CBS News [24] |
| Delta, UK, May 2021 | 60% (53%-66%) | Sheikh et.al [18] |

Based on the literature review, we known the VE to Delta variant is 69.89% (95%CI: 54.86%-84.92%) for Pfizer (BnT162b2); 74.47% (95%CI: 59.98%-89.95%) for Moderna (mRnA-1273); 62.67% (95%CI: 29.87%-95.47%) for J&J (Ad26.CoV2-S); 62.00% (95%CI: 56.64%-67.36%) for AZ (AZD1222). Therefore, the VE for the Omicron variant is estimated as 34.95% (95%CI: 27.43%-42.46%) for Pfizer (BnT162b2); 37.24% (95%CI: 29.99%-44.98%) for Moderna (mRnA-1273); 31.34% (95%CI: 14.94%-47.74%) for J&J (Ad26.CoV2-S); 31.00% (95%CI: 28.32%-33.68%) for AZ (AZD1222).

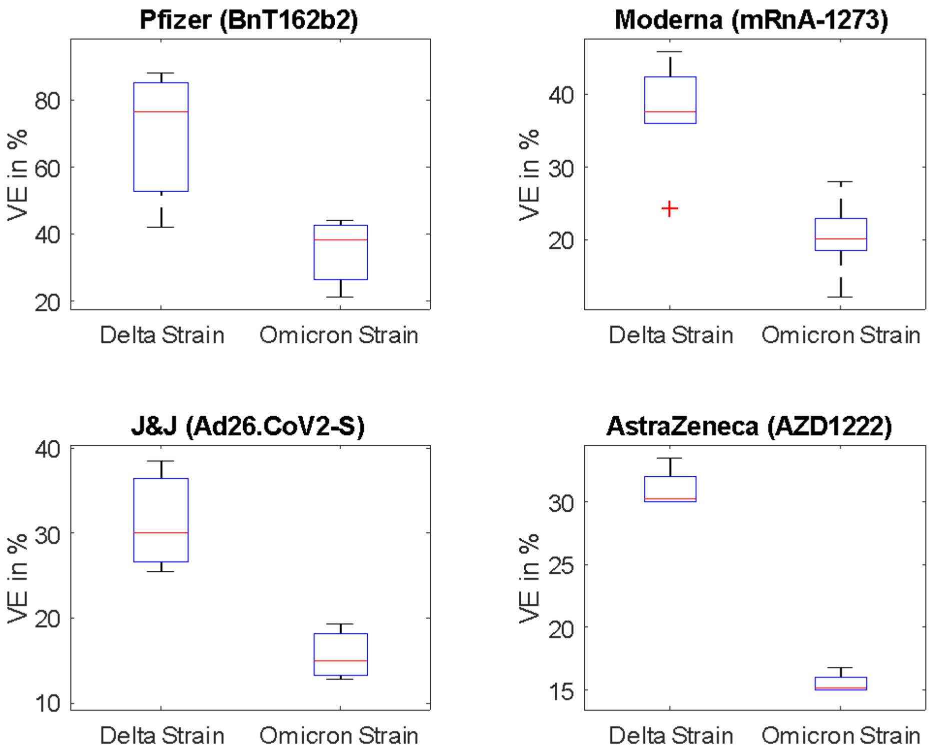

**Figure SI.5. VE to Delta and Omicron variants comparisons by the type of vaccines**

**The summarized data of the immunity level at studied LMICs**

There are significant differences in the level of immunity of the studied LMICs, due to the difference in types of vaccine, vaccination rate, and natural immunity coverage. We summarized the vaccination types and coverage rate for 25 studied LMICs from the Our World in Data group at the University Oxford [28], and estimated the nature immunity coverage rates based on WHO COVID-19 Dashboard [29] and several literatures with SARS-CoV-2 seroprevalence reported in LMICs [31-36]. The results show that only 2-30% COVID-19 cases were reported, therefore in this study we assumed that the actual prevalence is 8 times higher than the reported cases for the studied LMICs . Then the immunity efficiency is estimated based on the VE of the dominated vaccination type. The summarized data of immunity level at 25 studied LMICs are shown in Table SI.9.

**Table SI.9. The summarized data of the immunity at studied LMICs**

| LMICs | Dominated vaccination types (other vaccination types) | Vaccination rate (fully) | Nature Immunity Coverage | Estimated Immunity Efficiency |
|---|---|---|---|---|
| Bangladesh | AZ most (Moderna, Pfizer, Sinopharm/Beijing) | 22.74% | 7.58% | 31% |
| Nepal | AZ most (J&J, Pfizer, Sinopharm/Beijing) | 28.43% | 22.18% | 31% |
| Philippines | AZ most (J&J, Pfizer, Sinovac, Moderna, Sinopharm/Beijing) | 27.74% | 20.42% | 31% |
| Colombia | Pfizer most (AZ, Sinovac, J&J) | 49.1% | 39.64% | 34.95% |
| Egypt | AZ most (J&J, Pfizer, Sinopharm/Beijing) | 14.88% | 2.78% | 31% |
| Pakistan | AZ, CanSino, Sinopharm/Beijing | 23.23% | 4.58% | 31% |
| Paraguay | Pfizer most (AZ, covaxin, Sinovac, J&J) | 37.45% | 25.98% | 34.95% |
| Turkey | Pfizer most ( Sinovac) | 59.56% | 41.96% | 34.95% |
| Serbia | Pfizer most (AZ, Sinopharm/Beijing) | 45.65% | 58.2% | 34.95% |
| Bolivia | AZ most (J&J, Pfizer, Sinopharm/Beijing) | 35.41% | 36.72% | 31% |
| Argentina | AZ most (J&J, Moderna, Sinopharm/Beijing) | 66.36% | 45.34% | 31% |
| Uruguay | Pfizer most (AZ, Sinova) | 76.33% | 46% | 34.95% |
| Bhutan | AZ most (Moderna, Pfizer, Sinopharm/Beijing) | 72.36% | 2.72% | 31% |
| Indonesia | Moderna most (AZ, Pfizer, Snovac, Sinopharm/Beijing) | 35.83% | 13.32% | 37.24% |

| | | | | |
|---|---|---|---|---|
| Albania | Pfizer most (AZ, Sinovac) | 33.83% | 28.12% | 34.95% |
| Jordan | Pfizer most (AZ, Sinopharm/Beijing) | 36.77% | 28.24% | 34.95% |
| Panama | Pfizer most (AZ,) | 55.63% | 43.72% | 34.95% |
| Dominican | AZ most (Pfizer, Sinopharm/Beijing) | 51.31% | 29.84% | 31% |
| Ecuador | Pfizer most (AZ, Sinovac) | 64.3% | 23.6% | 34.95% |
| Peru | Pfizer most (AZ, Sinovac) | 55.69% | 26.88% | 34.95% |
| Jamaica | AZ most (J&J, Moderna, Pfizer) | 17.59% | 24.64% | 31% |
| Honduras | Moderna most (AZ, Pfizer, J&J) | 36% | 30.08% | 37.24% |
| Guatemala | Moderna most (AZ) | 22.88% | 27.2% | 37.24% |
| Costa Rica | Pfizer most (AZ) | 62.56% | 44.16% | 34.95% |
| El Salvador | AZ most (Pfizer, Sinovac) | 62.61% | 14.72% | 31% |

**Sensitivity analysis of the [illegible] of Omicron in LMICs**

With the immunity level data in LMICs, the value of $R_e$ can be calculated based on the equation (SI.5). We considered that the $R_0$of the Omicron variant as twice as the $R_0$of the Delta variant; the VE of the Omicron variant as 50% as the VE of the Delta variant [6]. And due to the differences in public health policies between LMICs, we didn't incorporate the intervention strategy and behavior in the estimated value of $R_e$ . In Figure SI.6 (a), we fixed the $R_0$of the Omicron variant as the baseline value, then allowing the VE of the Omicron variant as a free variable (between 30%-70% reduction of VE of the Delta variant). In Figure SI.6 (b), we fixed the VE of the Omicron variant as the baseline value, then allowing the $R_0$ of the Omicron variant as a free variable (between 1.5-2.25 * $R_0$ of the Delta variant).

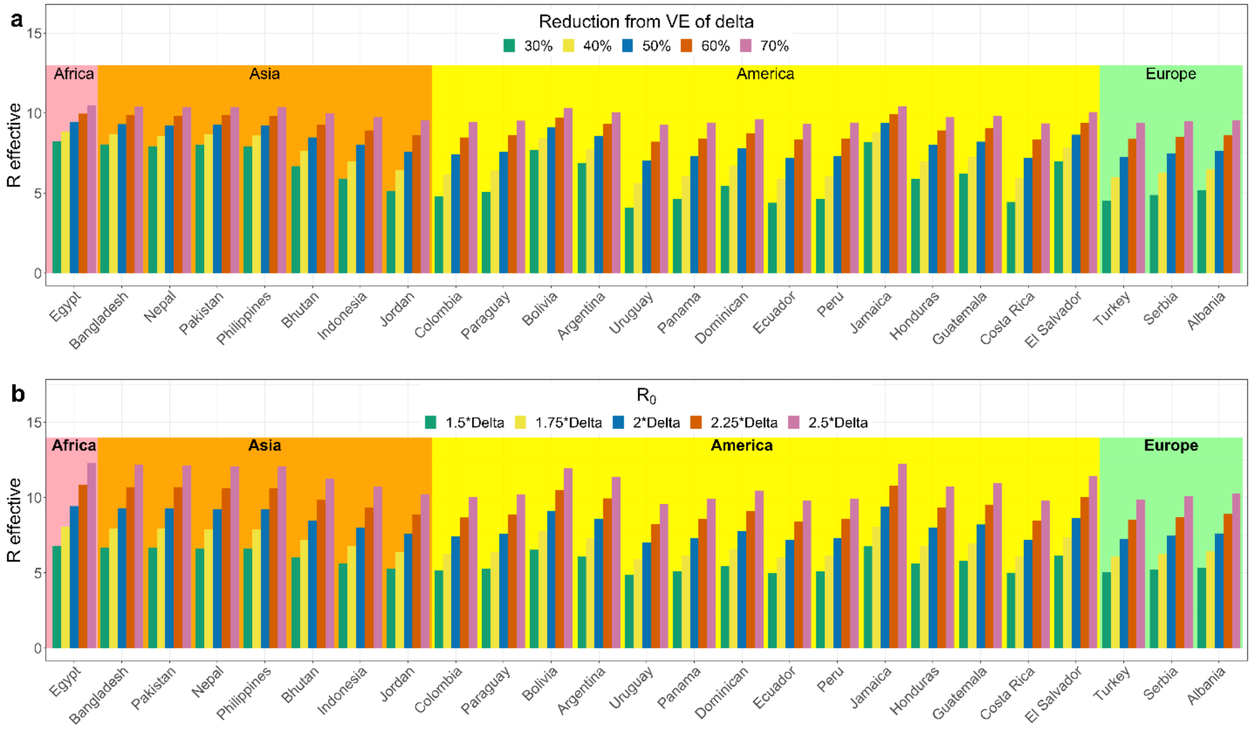


**Figure SI.6. The estimated value of $R_e$ for a given LMIC with $R_{0\_Omicron} = 2 * R_{0\_Delta}$(c) and $VE_{Omicron} = 50\% * VE_{Omicron}$ (d) as baselines, respectively.**

### Reference:


[1]: “Facebook Data for Good Travel Patterns.” n.d. Dataforgood.facebook.com. Accessed December 12, 2021. https://dataforgood.facebook.com/dfg/tools/travel-patterns.

[2]: Davis, Jessica T., Matteo Chinazzi, Nicola Perra, Kunpeng Mu, Marco Ajelli, Natalie E. Dean, Corrado Gioannini et al. "Cryptic transmission of SARS-CoV-2 and the first COVID-19 wave." Nature (2021): 1-9.

[3]: World Population Review. 2021. “Facebook Users by Country 2020.” Worldpopulationreview.com. 2021. https://worldpopulationreview.com/country-rankings/facebook-users-by-country.

[4]: “GISAID - HCov19 Variants.” n.d. Www.gisaid.org. https://www.gisaid.org/hcov19-variants/.

[5] Miller, Joel C., Anja C. Slim, and Erik M. Volz. "Edge-based compartmental modelling for infectious disease spread." Journal of the Royal Society Interface 9, no. 70 (2012): 890-906.

[6] Chen, Jiahui, Rui Wang, Nancy Benovich Gilby, and Guo-Wei Wei. "Omicron (B. 1.1. 529): Infectivity, vaccine breakthrough, and antibody resistance." arXiv preprint arXiv:2112.01318 (2021).

[7] Zhang, Meng, Jianpeng Xiao, Aiping Deng, Yingtao Zhang, Yali Zhuang, Ting Hu, Jiansen Li et al. "Transmission Dynamics of an Outbreak of the COVID-19 Delta Variant B. 1.617. 2—Guangdong Province, China, May–June 2021." China CDC Weekly 3, no. 27 (2021): 584-586.

[8] Qingfeng Shi, Xiaodong Gao, Bijie Hu. Research progress on characteristics, epidemiology and control measure of SARS-CoV-2 Delta VOC. Chin J Nosocomiol Vol.31, 2021

[9] “SPI-M-O: Summary of Further Modelling of Easing Restrictions -Roadmap Step 4 on 19.” n.d. Accessed October 24, 2021. https://assets.publishing.service.gov.uk/government/uploads/system/uploads/attachment_data/file/1001169/S1301_SPI-M-O_Summary_Roadmap_second_Step_4.2__1_.pdf.

[10] David Mackie, J.P. Morgan. Global vulnerabilities to the COVID-19 variant B.1.617.2. SUERF Policy Briefs No 110, June 2021

[11] Liu, Hengcong, Juanjuan Zhang, Jun Cai, Xiaowei Deng, Cheng Peng, Xinghui Chen, Juan Yang et al. "Herd immunity induced by COVID-19 vaccination programs to suppress epidemics caused by SARS-CoV-2 wild type and variants in China." medRxiv (2021).

[12] Head, Jennifer R., Kristin L. Andrejko, and Justin V. Remais. "Model-based assessment of SARS-CoV-2 Delta variant transmission dynamics within partially vaccinated K-12 school populations." medRxiv (2021).

[13] Hansena, Peter Reinhard. "Relative Contagiousness of Emerging Virus Variants: An Analysis of SARS-CoV-2 Alpha and Delta Variants." (2021).

[14] Mandal, Sandip, Nimalan Arinaminpathy, Balram Bhargava, and Samiran Panda. "Plausibility of a third wave of COVID-19 in India: A mathematical modelling based analysis." Indian J Med Res (2021).

[15] Nguyen, Trung, Mehnaz Adnan, Binh P. Nguyen, Joep de Ligt, Jemma L. Geoghegan, Richard Dean, Sarah Jefferies et al. "COVID-19 vaccine strategies for Aotearoa New Zealand: a mathematical modelling study." The Lancet Regional Health-Western Pacific 15 (2021): 100256.

[16] Alizon, Samuel, Stéphanie Haim-Boukobza, Vincent Foulongne, Laura Verdurme, Sabine Trombert-Paolantoni, Emmanuel Lecorche, Bénédicte Roquebert, and Mircea T. Sofonea. "Rapid spread of the SARS-CoV-2 Delta variant in some French regions, June 2021." Eurosurveillance 26, no. 28 (2021): 2100573.

[17] Campbell, Finlay, Brett Archer, Henry Laurenson-Schafer, Yuka Jinnai, Franck Konings, Neale Batra, Boris Pavlin et al. "Increased transmissibility and global spread of SARS-CoV-2 variants of concern as at June 2021." Eurosurveillance 26, no. 24 (2021): 2100509.

[18] Sheikh, Aziz, Jim McMenamin, Bob Taylor, and Chris Robertson. "SARS-CoV-2 Delta VOC in Scotland: demographics, risk of hospital admission, and vaccine effectiveness." The Lancet (2021).

[19] Lopez Bernal, Jamie, Nick Andrews, Charlotte Gower, Eileen Gallagher, Ruth Simmons, Simon Thelwall, Julia Stowe et al. "Effectiveness of Covid-19 vaccines against the B. 1.617. 2 (Delta) variant." N Engl J Med (2021): 585-594.

[20] Nasreen, Sharifa, Siyi He, Hannah Chung, Kevin A. Brown, Jonathan B. Gubbay, Sarah A. Buchan, Sarah E. Wilson et al. "Effectiveness of COVID-19 vaccines against variants of concern, Canada." Medrxiv (2021).

[21] Tang, Patrick, Mohammad Rubayet Hasan, Hiam Chemaitelly, Hadi M. Yassine, Fatiha Benslimane, Hebah A. Al Khatib, Sawsan AlMukdad et al. "BNT162b2 and mRNA-1273 COVID-19 vaccine effectiveness against the Delta (B. 1.617. 2) variant in Qatar." medRxiv (2021).

[22] Bajema, Kristina L., Rebecca M. Dahl, Mila M. Prill, Elissa Meites, Maria C. Rodriguez-Barradas, Vincent C. Marconi, David O. Beenhouwer et al. "Effectiveness of COVID-19 mRNA Vaccines Against COVID-19–Associated Hospitalization—Five Veterans Affairs Medical Centers, United States, February 1–August 6, 2021." Morbidity and Mortality Weekly Report 70, no. 37 (2021): 1294.

[23] Puranik, Arjun, Patrick J. Lenehan, Eli Silvert, Michiel JM Niesen, Juan Corchado-Garcia, John C. O'Horo, Abinash Virk et al. "Comparison of two highly-effective mRNA vaccines for COVID-19 during periods of Alpha and Delta variant prevalence." medRxiv (2021).

[24] CBS News. Delta variant of COVID-19 likely to become dominant U.S. strain, Gottlieb says - CBS News. https://www.cbsnews.com/news/covid-19-deltavarient- dominant-strain-likely/ (2021).

[25] Nanduri, Srinivas, Tamara Pilishvili, Gordana Derado, Minn Minn Soe, Philip Dollard, Hsiu Wu, Qunna Li et al. "Effectiveness of Pfizer-BioNTech and Moderna vaccines in preventing SARS-CoV-2 infection among nursing home residents before and during widespread circulation of the SARS-CoV-2 B. 1.617. 2 (Delta) variant—National Healthcare Safety Network, March 1–August 1, 2021." Morbidity and Mortality Weekly Report 70, no. 34 (2021): 1163.

[26] Polinski, Jennifer M., Andrew R. Weckstein, Michael Batech, Carly Kabelac, Tripthi Kamath, Raymond Harvey, Sid Jain, Jeremy A. Rassen, Najat Khan, and Sebastian Schneeweiss. "Effectiveness of the Single-Dose Ad26. COV2. S COVID Vaccine." medRxiv (2021).

[27] Barlow, Russell S., Lindsey Larson, and Kevin Jian. "Effectiveness of COVID-19 Vaccines Against SARS-CoV-2 Infection During a Delta Variant Epidemic Surge in Multnomah County, Oregon, July 2021." medRxiv (2021).

[28] Mathieu, E., Ritchie, H., Ortiz-Ospina, E. et al. A global database of COVID-19 vaccinations. Nat Hum Behav (2021). https://doi.org/10.1038/s41562-021-01122-8

[29] World Health Organization. 2021. "WHO COVID-19 Dashboard." Covid19.Who.int. World Health Organization. 2021. https://covid19.who.int/.

[30] Feehan, Dennis M., and Ayesha S. Mahmud. "Quantifying population contact patterns in the United States during the COVID-19 pandemic." Nature communications 12, no. 1 (2021): 1-9.

[31] Nisar, Muhammad Imran, Nadia Ansari, Mashal Amin, Farah Khalid, Aneeta Hotwani, Najeeb Rehman, Arjumand Rizvi et al. "Serial population based serosurvey of antibodies to SARS-CoV-2 in a low and high transmission area of Karachi, Pakistan." *MedRxiv* (2020).

[32] Bhuiyan, Taufiqur Rahman, Juan Dent Hulse, Sonia Hegde, Marjahan Akhtar, Taufiqul Islam, Zahid Hasan Khan, Ishtiakul Islam Khan et al. "SARS-CoV-2 seroprevalence in Chattogram, Bangladesh before a National Lockdown, March-April 2021." *medRxiv* (2021).

[33] Reyes-Vega, Mary F., M. Gabriela Soto-Cabezas, Fany Cárdenas, Kevin S. Martel, Andree Valle, Juan Valverde, Margot Vidal-Anzardo, María Elena Falcón, César V. Munayco, and Peru COVID-19 Working Group. "SARS-CoV-2 prevalence associated to low socioeconomic status and overcrowding in an LMIC megacity: A population-based seroepidemiological survey in Lima, Peru." *EClinicalMedicine* 34 (2021): 100801.

[34] Mugunga, Jean Claude, Kartik Tyagi, Daniel Bernal-Serrano, Nidia Correa, Matias Iberico, Frederick Kateera, Fernet Leandre, Megan Murray, Jean Christophe Dimitri Suffrin, and Bethany Hedt-Gauthier. "SARS-CoV-2 serosurveys in low-income and middle-income countries." *The Lancet* 397, no. 10272 (2021): 353-355.

[35] Rostami, Ali, Mahdi Sepidarkish, Mariska MG Leeflang, Seyed Mohammad Riahi, Malihe Nourollahpour Shiadeh, Sahar Esfandyari, Ali H. Mokdad, Peter J. Hotez, and Robin B. Gasser. "SARS-CoV-2 seroprevalence worldwide: a systematic review and meta-analysis." *Clinical Microbiology and Infection* 27, no. 3 (2021): 331-340.

[36] Rostami, Ali, Mahdi Sepidarkish, Aylar Fazlzadeh, Ali H. Mokdad, Aida Sattarnezhad, Sahar Esfandyari, Seyed Mohammad Riahi et al. "Update on SARS-CoV-2 seroprevalence: regional and worldwide." *Clinical Microbiology and Infection* 27, no. 12 (2021): 1762-1771.
